# Unchanged Early Diffusion Tensor Imaging Along Perivascular Space Index After Amyloid-Targeting Disease-Modifying Therapy in Alzheimer’s Disease: A Preliminary Study

**DOI:** 10.1101/2025.05.08.25327118

**Authors:** Tatsushi Oura, Hiroyuki Tatekawa, Akitoshi Takeda, Ayako Omori, Natsuko Atsukawa, Shu Matsushita, Daisuke Horiuchi, Hirotaka Takita, Taro Shimono, Daiju Ueda, Yoshiaki Itoh, Yukio Miki

## Abstract

**Purpose:** No longitudinal imaging biomarkers have been validated to capture early glymphatic changes during disease-modifying therapy (DMT) for Alzheimer’s disease (AD). This study investigated whether the diffusion tensor imaging along the perivascular space (DTI-ALPS) index can detect early treatment-related changes in participants with AD who initiate amyloid-targeting DMT.

**Methods:** Thirteen participants with AD (mean age, 72 years; 8 women) who initiated lecanemab therapy prospectively underwent DTI at baseline and three months. Projection and association fiber regions of interest, predefined in the HCP-1065 atlas, were inversely warped to the native space with vector-aware linear and non-linear registration, enabling a fully automated DTI-ALPS index calculation. Within-participant variances were obtained from 23 healthy volunteers in the OASIS dataset and used to set an equivalence margin of ±0.05 and determine a required sample size of 13. The primary end-point was equivalence of pre- and post-treatment DTI-ALPS indices (a two one-sided test; one-sided α = 0.05). Second, a paired *t*-test was used to assess the changes. The intraclass correlation coefficients (ICCs) of the DTI-ALPS indices were also evaluated in identical machine environments and between different environments.

**Results:** Baseline and three-month DTI-ALPS indices were 1.515 and 1.513, respectively. The mean change was 0.002 (90% confidence interval: –0.049, +0.045), entirely within the pre-specified margin; both one-sided *p*-values were < 0.05, confirming statistical equivalence. A paired *t*-test showed no significant difference (*p* = 0.94). Automated processing yielded perfect within-platform reproducibility (ICC = 1.00) and excellent cross-platform reliability (ICC = 0.99).

**Conclusion:** The DTI-ALPS index, an imaging metric associated with glymphatic activity, did not change during the first three months of lecanemab therapy. Although this finding suggests that glymphatic alterations may not be detectable early after treatment initiation, larger cohorts and longer follow-up periods are required to clarify the temporal relationship between DTI-ALPS index dynamics and therapeutic effects.

## Introduction

Alzheimer’s disease (AD) is characterized by the progressive accumulation of amyloid-β peptides in the brain parenchyma, and impairment of interstitial waste clearance via the glymphatic system is thought to be one of the causes (1,2). Recently approved disease-modifying (DM) monoclonal antibodies, such as lecanemab and donanemab, are expected to slow cognitive decline by improving amyloid β clearance (3–5). If amyloid is cleared more effectively, the glymphatic system should function better. However, direct in vivo evidence of such improvements remains scarce.

Diffusion-tensor imaging along the perivascular space (DTI-ALPS) index has emerged as a non-invasive surrogate marker that is associated with glymphatic activity (6,7). The DTI-ALPS index quantifies water diffusivity orthogonal to the major projection and association fibers abutting the medullary veins, and lower values are interpreted as reduced interstitial fluid movement. This index declines with normal aging and is significantly lower in patients with AD than in cognitively normal individuals (6,8). This reduction is thought to result from amyloid-β accumulation in the brain parenchyma, which impedes interstitial fluid movement along the glymphatic pathway.

However, to the best of our knowledge, no longitudinal imaging biomarkers have been validated to detect early glymphatic changes in patients with DM-treated AD. Clinically, the first three months of DM therapy (DMT) are critical because amyloid-related imaging abnormalities (ARIA), such as cerebral microhemorrhage or white matter edema, occur most frequently during this interval, which may presumably reflect the peak of amyloid clearance (9,10). Tracking the dynamics of the DTI-ALPS index in this window could provide an imaging biomarker of therapeutic efficacy and enable the early identification of responders. Notably, the DTI-ALPS index demonstrated excellent test-retest reliability (intraclass correlation coefficient [ICC] ≈ 0.95) in healthy adults, supporting its suitability for longitudinal monitoring (11).

Therefore, this study quantified the DTI-ALPS index in patients with AD before and three months after the initiation of amyloid-targeting DMT. We hypothesized that DMT would elevate the index, signifying improved glymphatic function, and thereby generate provisional reference values for the longitudinal assessment of future DM-treated cohorts.

## Materials and methods

This prospective study was approved by the ethics committee of our institution (IRB: 2023-136), and written informed consent was obtained from all participants. This study complied with the principles of the Declaration of Helsinki and was structured according to the Strengthening the Reporting of Observational Studies in Epidemiology (STROBE) guidelines (12).

### Study design and setting

This investigation comprised a two-step observational analysis.

1. Derivation: A publicly available Open Access Series of Imaging Studies (OASIS) dataset was used (http://oasis-brains.org/) (12). Among them, healthy volunteers (23 paired DTI data) who underwent two identical DTI acquisitions were used solely to characterize within-participant variability and to derive the sample size requirement and associated 90% confidence interval (CI) limits for the subsequent equivalence test.
2. Validation: Paired participants with pre- and post-treatment measurements (before and three months after the initiation of DMT) were recruited at our institution, and differences in the DTI-ALPS index were evaluated.

### Participants

Participants were prospectively and consecutively recruited between December 1, 2023, and February 28, 2025, if they met the following inclusion criteria: i) diagnosed with AD by neurologists; ii) underwent brain magnetic resonance imaging (MRI) as part of screening for DMT; and iii) were deemed eligible for DMT based on cerebrospinal fluid biomarkers or amyloid positron emission tomography findings, along with neurocognitive testing, and subsequently initiated lecanemab therapy. Only participants who provided written informed consent for this study and underwent DTI before treatment (baseline) and three months after initiation were included. Exclusion criteria included any case in which severe motion or other artifacts resulted in unacceptable image quality or failure of subsequent preprocessing. The selection flowchart is shown in Figure 1.

**Figure 1.**
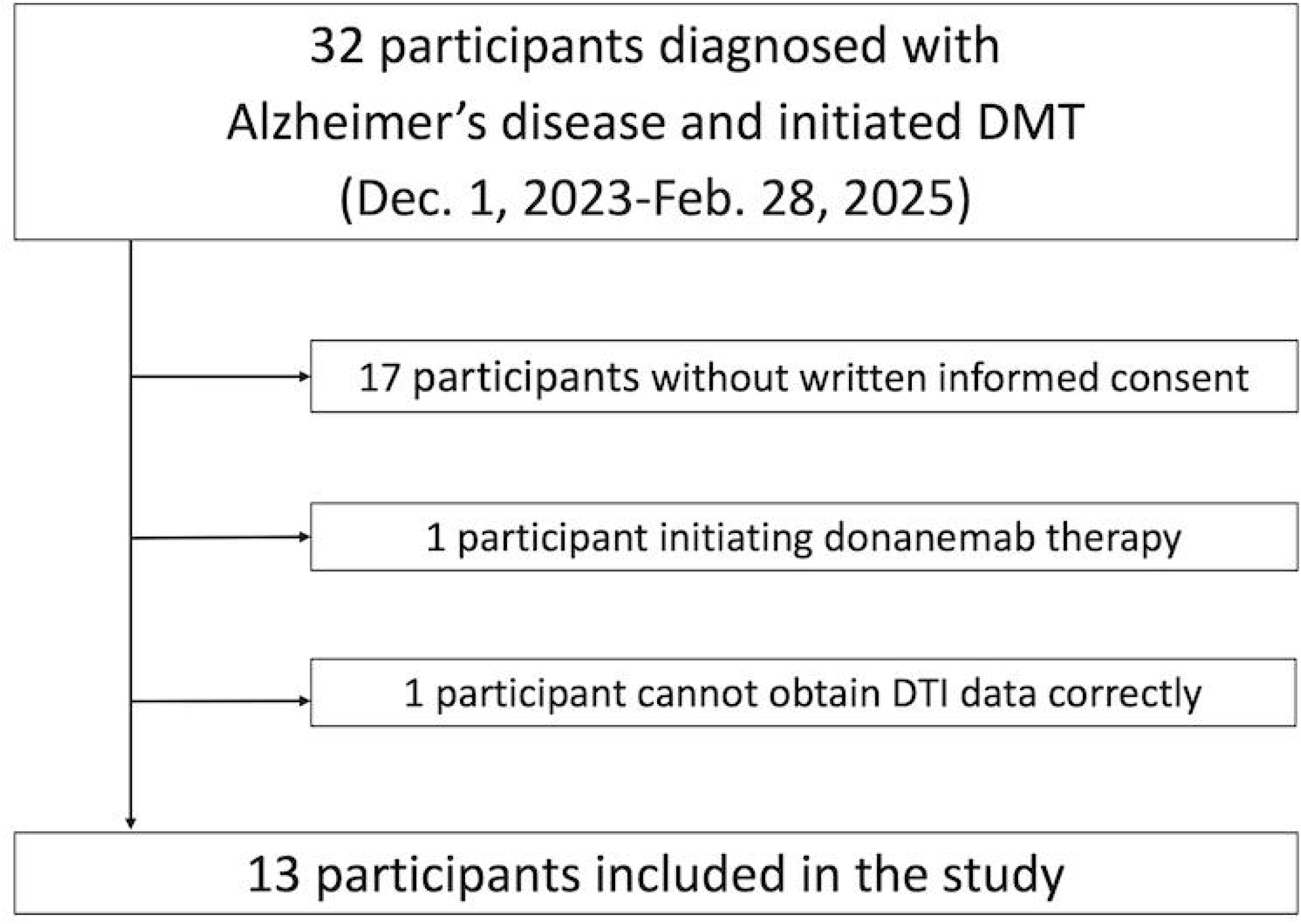
Selection flow chart. DMT, disease-modifying therapy; DTI, diffusion tensor imaging.

### MRI data acquisition

All examinations were performed on a 3 Tesla MRI system, Magnetom Vida (Siemens Healthineers, Erlangen, Germany; 20-channel head coil), with an identical single-shot echo-planar DTI protocol (TR/TE = 4500/81 ms; flip angle = 90°; voxel size = 2 × 2 × 2 mm^3^; b = 1000 s/mm^2^, 30 directions with one b0 volumes). Raw images were visually inspected by two radiologists (X.X. and Y.Y. with six- and 15-year radiology experiences, respectively); none were excluded. Baseline MRI was performed within three months before the first lecanemab dose, and follow-up MRI was performed exactly three months after treatment initiation.

### Measurements of DTI-ALPS index

Before analysis, every DTI data was first cleaned to reduce random noise, then stripped of Gibbs-ringing artifacts, corrected for motion- and eddy-current-related distortions, and finally adjusted for intensity bias, using the dedicated preprocessing tools available in MRtrix3.

Subsequently, the functions VECREG, FLIRT, and FNIRT, provided by the FMRIB software library (FSL), were used for DTI vector reorientation, linear registration, and non-linear registration, respectively. First, each participant’s DTI data was re-oriented using the previously described VECREG-based method (11). Second, linear (FLIRT) and non-linear (FNIRT) registration were applied to align the re-oriented fractional anisotropy (FA) maps to the FA map derived from the HCP1065 atlas provided in the FSL, with the purpose of generating the warp field required for subsequent inverse mapping. The HCP1065 atlas provides color-coded direction maps and other DTI-derived maps, thereby facilitating an intuitive definition of the region of interest (ROI). Spherical ROIs (φ = 5 mm) were placed on the bilateral projection and association fibers directly within the HCP1065 atlas space and then inversely warped to each participant’s re-oriented DTI space. The ROIs were visually inspected by two radiologists (X.X. and Y.Y.), and no manual adjustment of the ROI placement was required. Finally, the mean diffusivities along Dxx, Dyy, and Dzz within each warped ROI were extracted to compute the DTI-ALPS index using the conventional method (6). The processing overview and script are presented in Figure 2 and the Supplemental Data, respectively.

**Figure 2.**
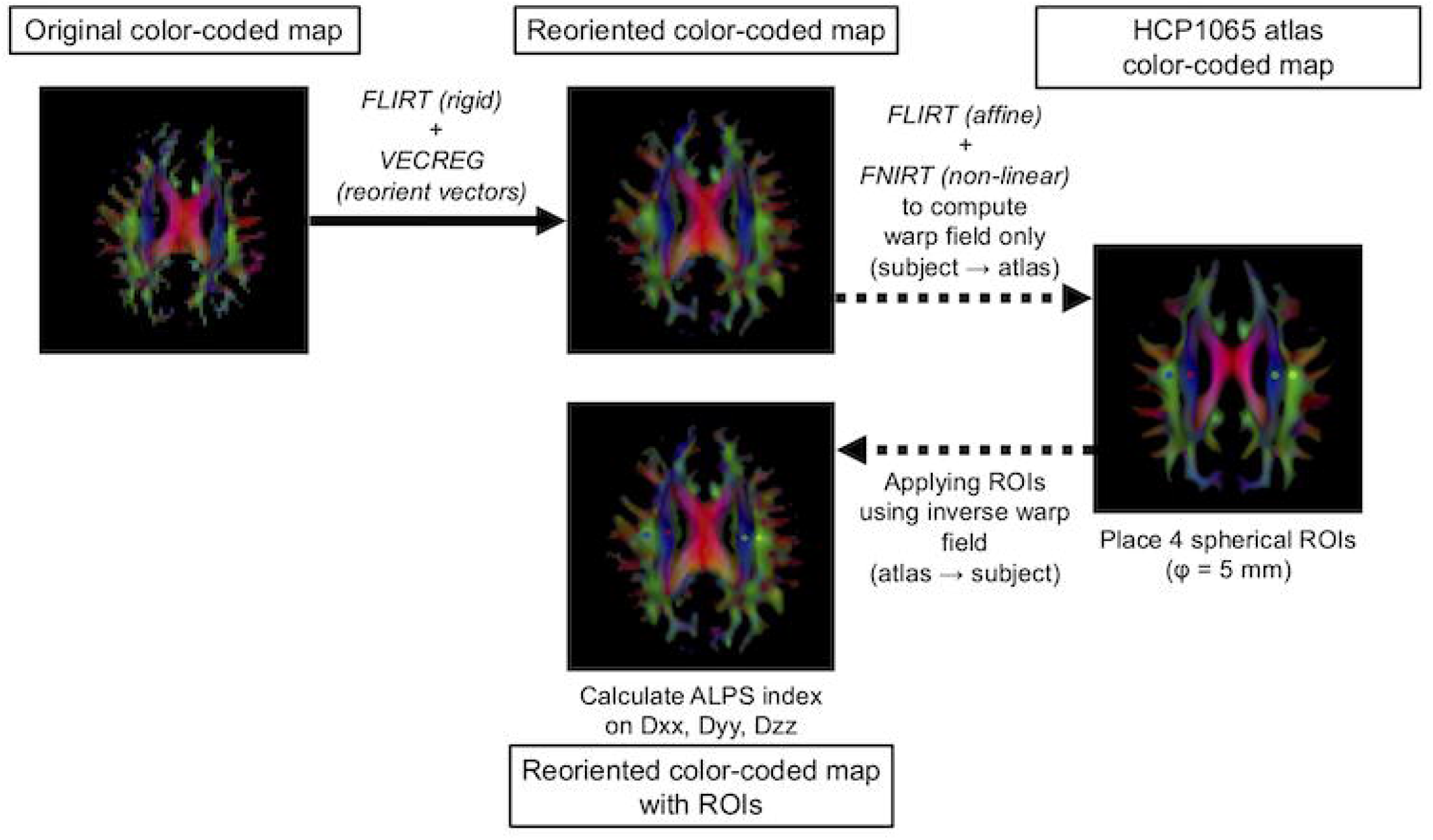
Workflow for automated DTI-ALPS index calculation. DTI data are visualized as color-coded direction maps for illustration purposes. 1) FA maps are rigidly aligned to the HCP1065 FA atlas using FLIRT, and tensor orientations are applied using VECREG to yield re-oriented maps (center-top). 2) The re-oriented FA maps are further aligned to the atlas with affine + non-linear registration (FLIRT → FNIRT) to generate a subject-to-atlas warp field (right). 3) In the atlas space, four spherical ROIs (diameter = 5 mm) are placed on the bilateral projection (blue) and association (green) fibers. 4) Using the inverse warp, these ROIs are mapped back to the re-oriented subject space and overlaid on the directional map (center-bottom). The mean diffusivities along Dxx, Dyy, and Dzz within each ROI are then extracted to compute the DTI-ALPS index. DTI, diffusion-tensor imaging; FA, fractional anisotropy; ROI, region of interest; DTI-ALPS, diffusion tensor imaging along the perivascular space.

### Sample-size calculation

This study primarily aimed to demonstrate equivalence between pre- and post-treatment measurements within a clinically relevant equivalence margin of ±0.05. This margin was selected on the basis of the observed within-participant standard deviation (0.07) derived from repeated DTI-ALPS index measurements in healthy volunteers (23 paired DTI scans) from the OASIS dataset, representing measurement reproducibility. Based on this within-participant standard deviation, a two one-sided test (TOST) with one-sided α = 0.05 and power = 0.80 indicated that a sample size of 13 paired participants was required. Raw data from the OASIS cohort and the MRI parameters are provided as Supplemental Data.

### Statistical analysis

Equivalence testing (primary objective): Equivalence of pre- and post-treatment measurements was assessed using a TOST with an equivalence margin (δ) of ±0.05 and one-sided α = 0.05. This margin was supported by the within-participant variability (standard deviation = 0.07) observed in the repeated DTI-ALPS index measurements from the healthy volunteer dataset (OASIS cohort, 23 paired measurements). Equivalence was concluded if both one-sided *p*-values were < 0.05, meaning the entire 90% CI of the mean difference between pre- and post-treatment DTI-ALPS indices lay entirely within the predefined ±0.05 margin.

Pre–post difference testing (secondary objective): To determine whether the treatment produced a statistically significant change, a paired *t*-test was applied. The ICCs of the DTI-ALPS index was calculated within an identical environment (Apple M1 max chip; FSL version 6.051) and between different environments (the previous environment and Apple M2 chip; FSL version 6.073).

All computations were performed in Python 3.11 with pandas 2.2 and SciPy 1.12.

## Results

Among the 32 participants who underwent DMT for AD, 13 met the inclusion criteria, and all participants were included based on the sample size power analysis. The mean age of the participants was 72 years, with a male:female ratio of 5:8. The mean Mini-Mental State Examination score was 24.6. The other demographic characteristics of the participants are summarized in Table 1.

**Table 1.**
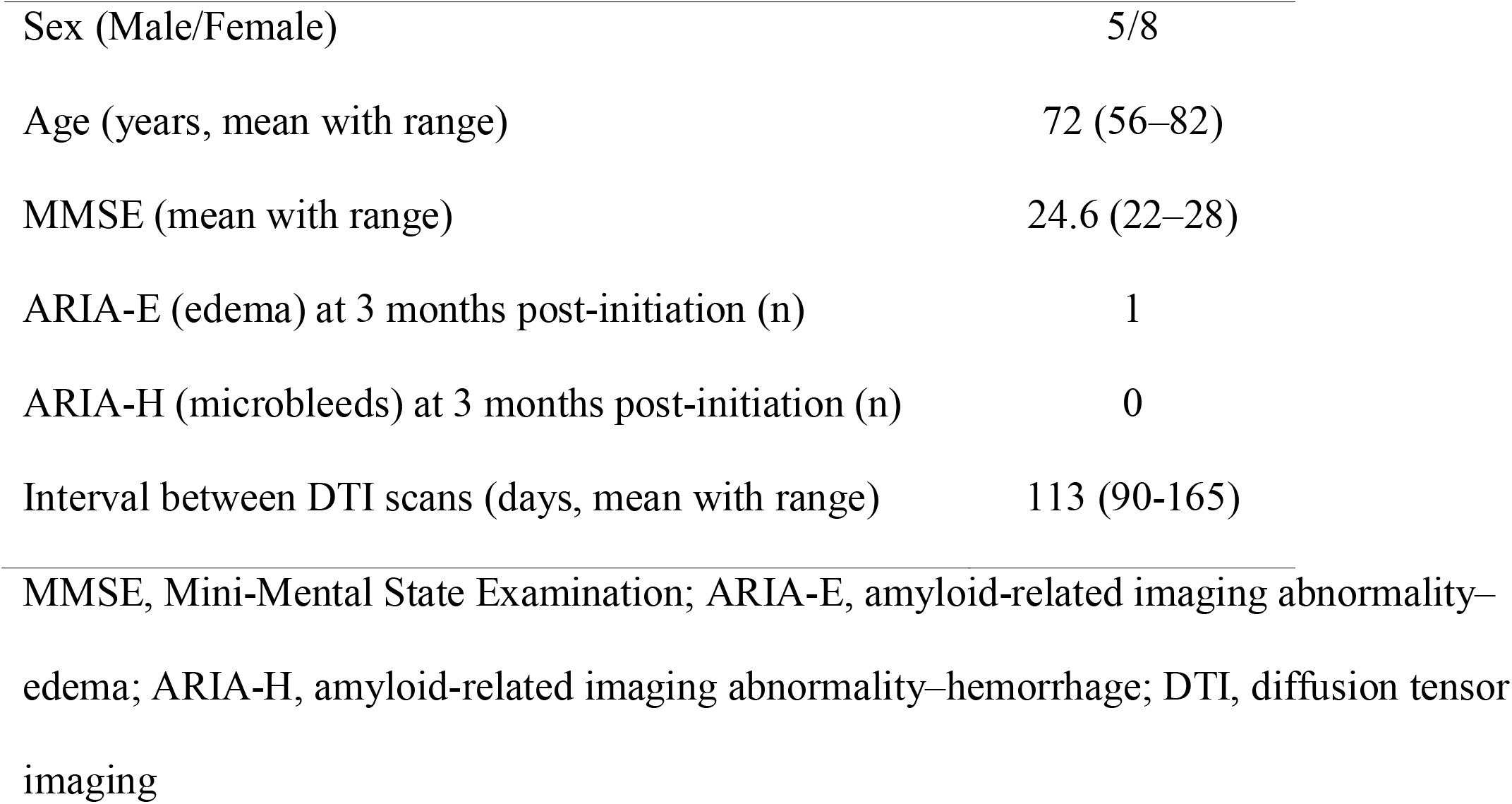
Characteristics and clinical data of the participants.

Primary outcome – equivalence test: The DTI-ALPS index averaged 1.515 ± 0.152 and 1.513 ± 0.161 at baseline and three months, respectively. The mean change in DTI-ALPS index from baseline to three months was δ = 0.002 units (90% CI = –0.049, +0.045). Because the entire CI lay within the prespecified equivalence margin of ±0.05, both one-sided TOST *p*-values were < 0.05 (*p*_*1*_ = 0.035; *p*_*2*_ = 0.047). These results demonstrate the statistical equivalence between the pre- and post-treatment DTI-ALPS indices.

Secondary outcome – paired *t*-test: A paired *t*-test showed no statistically significant differences between pre and post treatment (*t*_*12*_ = 0.08, two-tailed *p* = 0.94), supporting the finding that the treatment did not change the DTI-ALPS index within the first three months (Figure 3). Note, for the participant who occurred ARIA-E, the baseline DTI-ALPS index was 1.714, and that in three months was 1.727.

**Figure 3.**
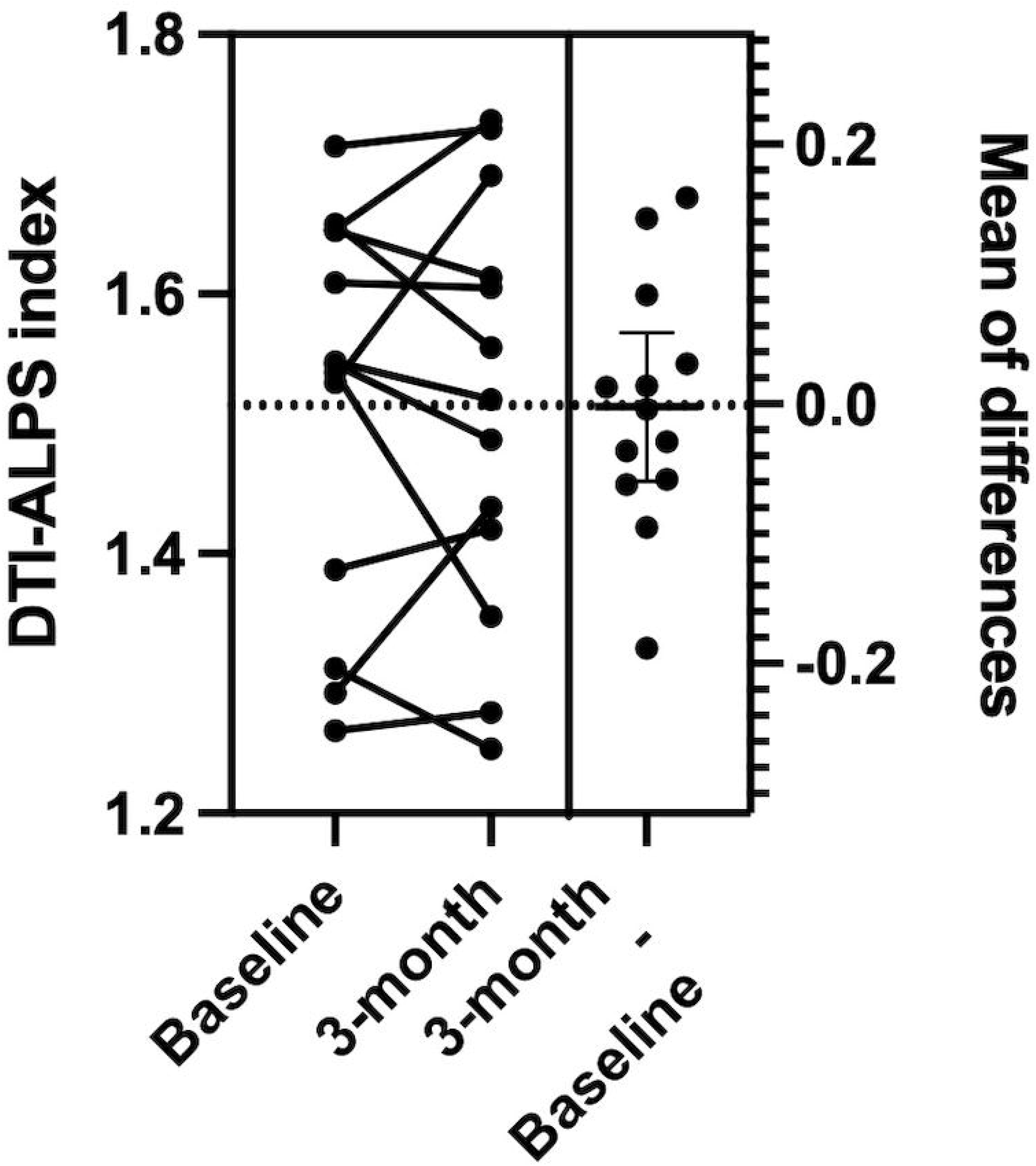
Dynamic change of DTI-ALPS index for pre- and post-DMT The paired t-test shows no statistically significant differences between pre- and post-DMT (*p* = 0.94). DTI-ALPS, diffusion tensor imaging along the perivascular space; DMT, disease-modifying therapy.

Repeated measurements of the automatic calculation of the DTI-ALPS index indicated perfect within-platform reproducibility (ICC = 1.00) and excellent cross-platform reliability (ICC = 0.99).

## Discussion

In this small but power-estimated cohort, the DTI-ALPS index showed no measurable improvement during the first three months of DMT. Although this is a small sample study, the findings can provide an initial benchmark for interpreting DTI-ALPS index in future studies evaluating the effect of DMT for AD.

The absence of early DTI-ALPS index improvement may reflect the chronic nature of amyloid deposition, which generally begins decades before clinical symptoms appear (14,15). DMT can reduce plaque burden and slow further cognitive worsening but does not restore lost function, likely reflecting the fact that neuronal damage and clearance system deficits have already been well established (3,5). This chronic nature of glymphatic dysfunction in AD implies that impaired waste clearance may continue even after amyloid removal, potentially allowing tau and other neurotoxic proteins to accumulate (2). Such observations likely reflect a multifactorial disease process that is not rapidly reversible during symptomatic stages.

Due to the small sample size in this study, multivariate or subgroup analyses were not feasible. However, future investigations should examine the relationship between the DTI-ALPS index changes and several potentially important factors. For instance, age-related differences in the glymphatic function response may be critical, given that glymphatic clearance reportedly declines significantly with advancing age (16). Sex differences might also influence glymphatic system activity, as recent imaging data have indicated that women generally exhibit higher DTI-ALPS indices than men, although this advantage may diminish significantly after menopause (17). Moreover, baseline cognitive function and the presence of the APOE ε4 genotype, both known to significantly influence AD progression, should also be explored as possible modifiers of glymphatic response. APOE ε4 carriers, in particular, exhibit both increased risk of ARIA and possibly greater cognitive benefits from DMT, emphasizing the importance of genotype considerations (18). Furthermore, evaluating the association between changes in the DTI-ALPS index and ARIA incidence could enable this biomarker to serve as a predictive tool for ARIA risk, further enhancing its clinical utility (19). Finally, because this study evaluated only the initial three-month changes in the DTI-ALPS index, identifying the characteristics of responders versus non-responders to amyloid-targeting therapies in longitudinal cohorts could provide crucial insights for personalized treatment strategies.

Notably, the automatic calculation of DTI-ALPS index presented in this study indicated almost perfect reproducibility (ICC = 0.99–1.00). Although the optimal method to quantify glymphatic function using the DTI-ALPS index is still debatable, our pipeline eliminates the need for manual ROI drawing in the brain for each participant with accounting for vector information (7,11). As it is based on non-linear registration (FNIRT), the same framework can also be applied to align and analyze other brain structures precisely from the DTI atlas. However, FNIRT may fail in brains with pronounced morphological abnormalities; therefore, automatically generated ROIs should always be inspected in the native space to confirm the correct placement.

This study had some limitations. First, the sample size was small, which prevented reliable subgroup or multivariate analyses. Larger investigations with long-term studies are needed to confirm the results. Second, only participants treated with lecanemab were included in this study. As other DMT for AD, such as donanemab, are also used, future research should compare these agents to determine whether the findings hold across therapies. Finally, although APOE ε4 genotype data were not available in the current study, future research should examine the genotype as it significantly influences AD progression and ARIA risk.

In conclusion, although DMT promotes β-amyloid clearance, the DTI-ALPS index, which is an imaging metric associated with glymphatic activity, did not change during the first three months of lecanemab therapy. Although this finding suggests that glymphatic alterations may not be detectable early after treatment initiation, larger cohorts and longer follow-up periods are required to clarify the temporal relationship between DTI-ALPS index dynamics and therapeutic effects. The automatic calculation of the DTI-ALPS index presented in this study indicated almost perfect reproducibility and would be useful for future studies to calculate the DTI-ALPS index.

## Supporting information

Script

ROI data

## Data Availability

All data produced in the present study are available upon reasonable request to the authors

## Acknowledgment

During the preparation of this study, the authors used ChatGPT, which is based on the GPT-4o architecture, to improve readability and language. After using this tool, the authors reviewed and edited the content as needed and take full responsibility for the content of the publication.

