## Supplementary material for "Unchanged Early Diffusion Tensor Imaging Along Perivascular Space Index After Amyloid-Targeting Disease-Modifying Therapy in Alzheimer’s Disease: A Preliminary Study": Script: Automatic_ALPS_calculator.sh

```
#!/bin/bash

## Please prepare dti.bval	dti.bvec	dti.json	dti.nii.gz files
## Please create acqparams.txt and index.txt

###############################################################################
# WARNING
# -------
# 1. As of FSL 6.0.7, the utility ‘vecreg’ is no longer included in the official
#    FSL distribution.  Install it manually or replace it with an equivalent
#    tool (e.g., ANTs ReorientTensorImage or MRtrix3 transformcalc) before
#    running this pipeline.
#
# 2. Ensure that the environment variable $FSLDIR is exported
#    and point to your FSL installation directory.  
#    The script will abort if it is undefined.
###############################################################################

#########################################
## Step 1: Brain extraction from b=0
#########################################
# Extract the first b=0 volume
#fslroi dti.nii.gz b0 0 1

# Brain extraction to create mask
#bet b0 b0_brain -m -R -f 0.25 -g 0.05  # Generates b0_brain_mask.nii.gz

#########################################
## Step 2: Eddy current correction
#########################################
#eddy \
#  --imain=dti.nii.gz \
#  --mask=b0_brain_mask.nii.gz \
#  --acqp=acqparams.txt \
#  --index=index.txt \
#  --bvecs=dti.bvec \
#  --bvals=dti.bval \
#  --out=dti_eddy_corrected \
#  --slm=linear

#########################################
## Step 3: Tensor fitting and FA map generation
#########################################
#dtifit \
#  --data=dti_eddy_corrected.nii.gz \
#  --out=dti \
#  --mask=b0_brain_mask.nii.gz \
#  --bvecs=dti_eddy_corrected.eddy_rotated_bvecs \
#  --bvals=dti.bval \
#  --save_tensor

#########################################
## When use MRtrix3
#########################################
## Step 1-2: Prepare preprocessed images (existing code reused)
mrconvert dti.nii.gz dti.mif -fslgrad dti.bvec dti.bval -datatype float32  # MRtrix3
dwidenoise dti.mif dti_den.mif -noise dti_noise.mif  # MRtrix3
mrdegibbs dti_den.mif dti_den_unr.mif -axes 0,1  # MRtrix3
dwifslpreproc dti_den_unr.mif dti_den_unr_preproc.mif -pe_dir AP -rpe_none -eddy_options "--slm=linear"  # MRtrix3
dwibiascorrect ants dti_den_unr_preproc.mif dti_den_unr_preproc_unbiased.mif -bias bias.mif  # MRtrix3
dwi2mask dti_den_unr_preproc_unbiased.mif mask_den_unr_preproc_unbiased.nii.gz  # MRtrix3
mrconvert dti_den_unr_preproc_unbiased.mif dti_den_unr_preproc_unbiased.nii.gz -export_grad_fsl dti_den_unr_preproc_unbiased.bvec dti_den_unr_preproc_unbiased.bval  # MRtrix3

## Step 3: Create diffusion tensor and maps
# Generate tensor and diffusion metrics
dtifit --bvals=dti_den_unr_preproc_unbiased.bval --bvecs=dti_den_unr_preproc_unbiased.bvec --data=dti_den_unr_preproc_unbiased.nii.gz --mask=mask_den_unr_preproc_unbiased.nii.gz --out=dti --save_tensor  # FSL

#########################################
## Step 4: Linear registration to FSL_HCP1065_FA_1mm
#########################################
ATLAS_FA="$FSLDIR/data/standard/FSL_HCP1065_FA_1mm.nii.gz"

# Rigid-body (6 DOF) alignment of FA map to the standard atlas
flirt -ref $ATLAS_FA -in dti_FA.nii.gz -omat fa2HCP1065_rigid.mat -dof 6 \
      -searchrx -90 90 -searchry -90 90 -searchrz -90 90

# Apply linear transformation to FA map
flirt -ref $ATLAS_FA -in dti_FA.nii.gz -applyxfm -init fa2HCP1065_rigid.mat -out dti_FA_ATLAS.nii.gz

#########################################
## Step 5: Tensor and eigenvector reorientation using vecreg
#########################################
# Requires vecreg to be installed separately (part of FSL or external)
# Reorient tensor image using linear transform
vecreg -i dti_tensor.nii.gz -o dti_ATLAS_tensor.nii.gz -r $ATLAS_FA -t fa2HCP1065_rigid.mat

# Reorient principal eigenvector image (V1)
vecreg -i dti_V1.nii.gz -o dti_ATLAS_V1.nii.gz -r $ATLAS_FA -t fa2HCP1065_rigid.mat

#########################################
## Step 6: Extract individual tensor components and visualize directionality
#########################################

# Extract transformed tensor components from reoriented tensor image
# Dxx (component 0)
#fslroi dti_ATLAS_tensor.nii.gz dti_ATLAS_Dxx.nii.gz 0 1
# Dyy (component 3)
#fslroi dti_ATLAS_tensor.nii.gz dti_ATLAS_Dyy.nii.gz 3 1
# Dzz (component 5)
#fslroi dti_ATLAS_tensor.nii.gz dti_ATLAS_Dzz.nii.gz 5 1

# Create a directionally encoded color FA (DEC-FA) image by multiplying V1 with FA
#fslmaths dti_ATLAS_V1.nii.gz -mul dti_FA_ATLAS.nii.gz dti_ATLAS_DEC.nii.gz
# Visualize the DEC-FA image overlaid on FA using fsleyes

# afni can also create Color maps
3dcalc -prefix DTIout_ATLAS.nii.gz -a 'dti_ATLAS_V1.nii.gz[0..2]' -c 'dti_FA_ATLAS.nii.gz' -expr 'c*step(c-0.2)*255*abs(a)'
3dThreetoRGB -prefix DTI_ATLAS_ColorMap.nii.gz -anat DTIout_ATLAS.nii.gz


#########################################
## Step 7: Nonlinear registration to standard space
#########################################
flirt -in dti_FA_ATLAS.nii.gz \
      -ref $ATLAS_FA \
      -omat fa2HCP1065_affine.mat \
      -out dti_FA_flirt.nii.gz

fnirt --in=dti_FA_ATLAS.nii.gz \
      --aff=fa2HCP1065_affine.mat \
      --ref=$ATLAS_FA \
      --iout=dti_FA_warped_to_HCP1065.nii.gz \
      --cout=dti_to_HCP1065_warp.nii.gz \
      --config=$FSLDIR/etc/flirtsch/FA_2_FMRIB58_1mm.cnf

#########################################
## Step 8: Inverse warp to transform ROI from standard to native space
#########################################
invwarp --warp=dti_to_HCP1065_warp.nii.gz \
        --ref=dti_FA_ATLAS.nii.gz \
        --out=HCP1065_to_dti_warp.nii.gz

# ROI label definitions:
# Please change the path of ROI data (FSL_HCP1065_FA_ROI.nii.gz)
# 1 = right CR, 2 = left CR, 3 = right SLF, 4 = left SLF
ATLAS_ROI=Path/to/FSL_HCP1065_FA_ROI.nii.gz # Please define it
applywarp --in=$ATLAS_ROI \
          --ref=dti_FA_ATLAS.nii.gz \
          --warp=HCP1065_to_dti_warp.nii.gz \
          --interp=nn \
          --out=ROI_in_dti_FA_ATLAS_space.nii.gz

#########################################
# Please review whether ROI data (ROI_in_dti_FA_ATLAS_space.nii.gz)
# is appropriately located in the DTI direction map (dti_FA_ATLAS.nii.gz dti_ATLAS_DEC.nii.gz or DTI_ATLAS_ColorMap.nii.gz).
# Then, please calculate ALPS index from dti_ATLAS_Dxx.nii.gz, dti_ATLAS_Dyy.nii.gz, and dti_ATLAS_Dzz.nii.gz
# or dti_ATLAS_V1.nii.gz using ROI_in_dti_FA_ATLAS_space.nii.gz
#########################################
```
