## Supplemental data for "Unchanged Early Diffusion Tensor Imaging Along Perivascular Space Index After Amyloid-Targeting Disease-Modifying Therapy in Alzheimer’s Disease: A Preliminary Study"

| **Supplemental Table 1.** DTI-ALPS index for normal control of OASIS dataset | |
| --- | --- |
| Age (years) | 66 ± 16.9 |
| Sex (Females/males) | 10/13 |
| MMSE | 29.64 ± 0.79 |
| Interval (days) | 709 ±328 |
| Test | 1.567 ± 0.184 |
| Retest | 1.585 ± 0.201 |
| Difference | -0.018 ± 0.07 |
| Values are presented as mean ± standard deviation, except for Sex, which is shown as number of females/males. | |

**DTI parameters of OASIS dataset**

All MRI data from the OASIS-3 database were acquired using a 3 Tesla scanner (Siemens Healthineers, Erlangen, Germany) with an identical single‑shot echo‑planar DTI protocol (TR/TE = 11000/87 ms; flip angle = 90°; voxel size = 2.5 × 2.5 × 2.5 mm³; b = 1000 s/mm², 64 directions with one b0 volume). Detailed information is available on the OASIS website.
